# Selection without Collider Stratification can Bias Instrumental Variable Analyses in the Presence of Causal Effect Heterogeneity

**DOI:** 10.64898/2026.09.27.26364122

**Authors:** Apostolos Gkatzionis, Fernando Pires Hartwig, Kate Tilling

## Abstract

Selection bias is a common concern in causal inference. Two mechanisms can give rise to selection bias: collider bias or stratifying on an effect modifier of the exposure-outcome causal effect. Research into the impact of selection bias on instrumental variable (IV) analyses has focused primarily on collider bias. Here, we show that IV estimates can be biased if selection is an effect modifier (or caused by an effect modifier) of the exposure-outcome effect. In a simulation study, we investigate the size and direction of the bias when estimating effects on the mean difference or odds ratio scale. Bias can also occur due to heterogeneity in the instrument-exposure relationship, although its magnitude is typically smaller. Finally, we discuss methods to deal with this bias in practice, including inverse probability weighting (IPW).

## 1. Introduction

Instrumental variable (IV) analysis assesses the causal effect of an exposure on an outcome in the presence of unmeasured confounding. The “instrument” is a variable that satisfies three assumptions: (IV1) it is robustly associated with the exposure, (IV2) it does not share common causes with the outcome, (IV3) it only affects the outcome through the exposure. Though less susceptible to confounding bias, IV analysis is still vulnerable to other biases, including selection bias. Papers investigating the impact of selection bias on IV analyses [1-3] have focused on bias due to conditioning on one or more levels of a collider [4]. However, selection bias does not always arise from collider stratification [5, 6]. Here, we describe an alternative mechanism and illustrate it in a simulation study.

## 2. Selection Bias, Collider Bias and Heterogeneity

Before focusing on IV analyses, we review selection bias in general. Let *X* be an exposure, *Y* an outcome and suppose that our objective is to estimate the *X*-*Y* average causal effect (ACE) in a target population. For simplicity, we assume in this section that other biases (e.g. confounding) have been appropriately dealt with and focus on selection bias.

To estimate the ACE, suppose we have a sample from the target population. Let *S* be a selection indicator: for individual *i*, if *S*_*i*_ = 1, that individual is included in the sample and we observe (*X*_*i*_, *Y*_*i*_), while if *S*_*i*_ = 0, the individual is not included in the sample and (at least one of) their *X*_l_, *Y*_l_ are unobserved. With the available data, a standard regression analysis can be used to estimate the ACE in the selected sample (conditional on *S* = 1). Selection bias occurs when the sample estimate of the ACE is not an (asymptotically) unbiased estimate of the true population-level effect [5].

Structurally, selection bias often occurs as collider bias. A collider is a variable that is causally downstream of two other variables; conditioning on a collider can distort the association between the variables causing it (Figure 1a). In practice, selection into a study often functions as a collider: if the exposure (or a cause of it) and the outcome (or a cause of it) affect individuals’ likelihood of being included in the sample, ACE estimates obtained in that sample may be biased by selection [4].

**Figure 1.**
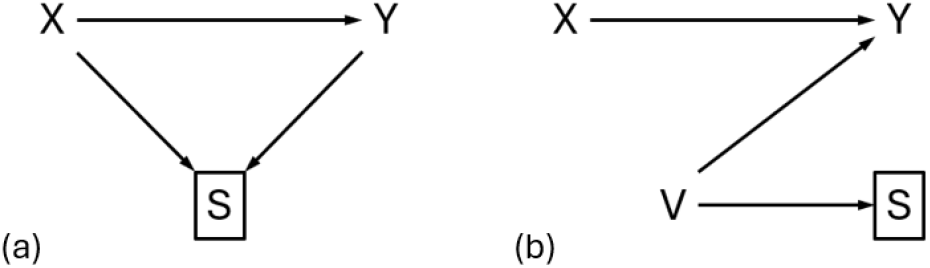
Causal diagrams illustrating selection bias due to conditioning on a collider (a) and due to heterogeneity (b).

However, selection bias can be induced even in the absence of collider bias. Consider the directed acyclic graph (DAG) of Figure 1b and assume that variable *V* modifies the exposure-outcome effect. Here, collider bias is not induced as there is only one cause of

*S*. However, selection bias may still be induced: *X* and *V* are common causes of *Y*, so they could be interacting in their effects on it. In this case, the ACE is effectively averaged across levels of *V*. As the variable *V* affects selection, its distribution in the selected sample will differ from that in the target population. Therefore, a sample estimate of the ACE will average over a different distribution of *V* than that in the target population, resulting in bias. This has been called selection bias due to heterogeneity [6] or Type 2 selection bias [5]. It is viewed as affecting the generalizability of a study’s results, while collider bias affects the study’s internal validity [5]. This bias does not occur under the sharp causal null hypothesis, as in this case there is no heterogeneity.

## 3. Selection Bias in Instrumental Variable Analyses

The impact of selection bias on IV studies has been investigated by several authors [1-3]. These papers have focused on collider bias, although Hughes et al. [2] acknowledged that selection bias can occur in the absence of collider bias. Here, we focus on bias due to heterogeneity; we briefly discuss how collider bias affects IV analyses in Supplementary Section 1.

Consider the DAG of Figure 2a, where variable V affects selection and the outcome. In this diagram, V is the only cause of S, therefore S is not a collider and conditioning on it will not induce collider bias in the IV analysis. However, as X and V are both causes of Y, V may modify the effect of X on Y. In practice, V may be unobserved; even if observed, a researcher performing IV analysis to estimate the ACE may choose not to adjust for it (adjusting for V would estimate a conditional effect). This can induce bias similar to Figure 1b: the distribution of V in the selected sample is different to that in the target population, therefore an IV estimate of the ACE in the selected sample may be a biased estimate of the population-level effect.

**Figure 2.**
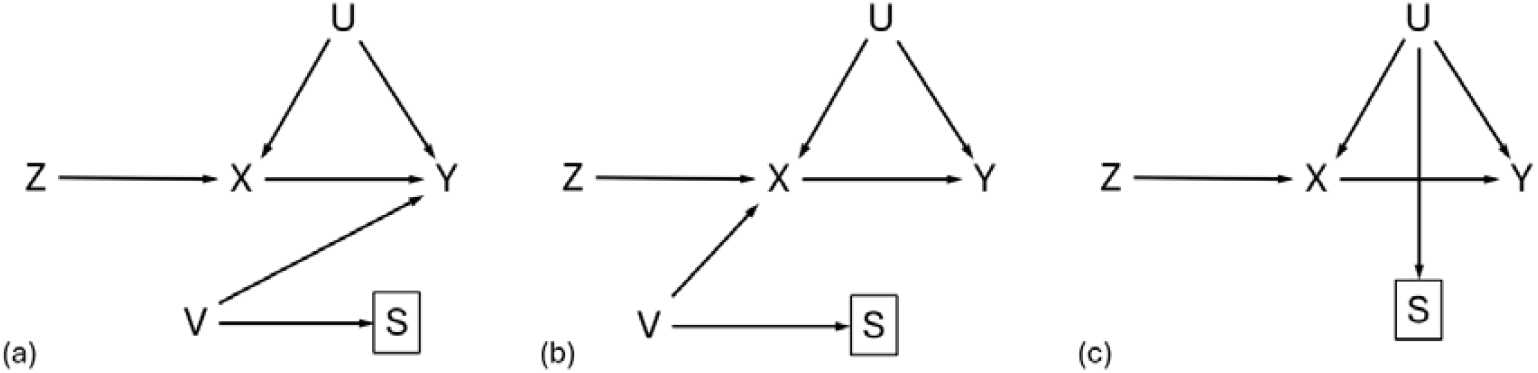
DAGs representing IV analyses where selection bias can be induced due to causal effect heterogeneity.

Similarly in Figure 2b, V can be an effect modifier for the effect of the instrument Z on exposure X. This can induce bias in a Z-X regression, which is the first step to running the IV analysis, and ultimately bias IV estimates. Finally, in Figure 2c, the confounder U can be an effect modifier for either the Z-X or the X-Y effect, so selection bias due to heterogeneity may occur.

## 4. Simulation Study

We conducted a simulation study to demonstrate selection bias due to heterogeneity in IV analyses. The simulation design is detailed in Supplementary Section 2. Briefly, we investigated two scenarios, corresponding to the DAGs of Figures 2a and 2b. We considered a normally distributed exposure and a continuous or binary outcome. We performed IV analysis using the two-stage least squares (TSLS) method for continuous *Y* and the two-stage residual inclusion (TSRI) method for binary *Y*. Our primary performance measure was the bias in exposure-outcome effect estimates obtained from a “complete-records” analysis using only the selected sample. Secondarily, we computed model-based standard errors and assessed the empirical coverage of 95% confidence intervals from the TSLS and TSRI fits.

Results are reported in Table 1. In simulations with heterogeneity in the *X*-*Y* effect, the impact of selection bias on IV estimates was clear for both continuous and binary outcomes. Stronger selection effects (*δ*_*VS*_) and stronger effect modification (*β*_*XVY*_ or, *β*_*ZVX*_) resulted in more bias and worse empirical coverage. When, *β*_*XVY*_ =0 or, *β*_*ZVX*_ = 0, no bias was induced.

**Table 1.** Bias, model-based standard error (“Se”) and empirical coverage (“Cov”) of TSLS/TSRI estimates for various parameter values in our simulation study: *δ*_*XY*_ represents the exposure-outcome causal effect, *β*_*VS*_ the effect of the effect modifier *V* on selection, and *β*_*XVY*_ (*β*_*ZVX*_) the strength of interaction between *X* and *V* in their effects on *Y* (or *Z* and *V* in their effects on *X*).

| X – Y heterogeneity |  |  |  |  |  |  |  |  |  |  |  |  |  |  |  |  |  |
| --- | --- | --- | --- | --- | --- | --- | --- | --- | --- | --- | --- | --- | --- | --- | --- | --- | --- |
| Y | $\beta_{XY}$ | $\delta_{VS}$ | $\beta_{XVY} = -0.5$ | | | $\beta_{XVY} = -0.25$ | | | $\beta_{XVY} = 0$ | | | $\beta_{XVY} = 0.25$ | | | $\beta_{XVY} = 0.5$ | | |
|  |  |  | Bias | Se | Cov | Bias | Se | Cov | Bias | Se | Cov | Bias | Se | Cov | Bias | Se | Cov |
| Cont | 0 | 0.2 | -0.064 | 0.077 | 0.889 | -0.034 | 0.072 | 0.935 | 0.001 | 0.071 | 0.962 | 0.029 | 0.072 | 0.931 | 0.062 | 0.077 | 0.869 |
|  |  | 0.5 | -0.149 | 0.075 | 0.503 | -0.071 | 0.071 | 0.853 | -0.001 | 0.070 | 0.950 | 0.070 | 0.071 | 0.812 | 0.145 | 0.075 | 0.501 |
|  |  | 1 | -0.253 | 0.072 | 0.059 | -0.126 | 0.068 | 0.535 | -0.000 | 0.067 | 0.953 | 0.125 | 0.068 | 0.531 | 0.252 | 0.072 | 0.090 |
|  | 0.5 | 0.2 | -0.061 | 0.077 | 0.892 | -0.030 | 0.072 | 0.939 | -0.003 | 0.071 | 0.948 | 0.027 | 0.072 | 0.915 | 0.055 | 0.077 | 0.862 |
|  |  | 0.5 | -0.144 | 0.075 | 0.523 | -0.072 | 0.071 | 0.840 | -0.002 | 0.070 | 0.952 | 0.071 | 0.071 | 0.814 | 0.146 | 0.075 | 0.494 |
|  |  | 1 | -0.253 | 0.072 | 0.053 | -0.129 | 0.068 | 0.547 | 0.001 | 0.067 | 0.940 | 0.126 | 0.068 | 0.529 | 0.244 | 0.072 | 0.107 |
| Bin | 0 | 0.2 | -0.036 | 0.087 | 0.925 | -0.019 | 0.088 | 0.945 | -0.004 | 0.088 | 0.957 | 0.015 | 0.087 | 0.949 | 0.037 | 0.087 | 0.925 |
|  |  | 0.5 | -0.087 | 0.086 | 0.847 | -0.042 | 0.087 | 0.931 | -0.006 | 0.088 | 0.949 | 0.042 | 0.088 | 0.918 | 0.086 | 0.088 | 0.834 |
|  |  | 1 | -0.151 | 0.085 | 0.580 | -0.078 | 0.086 | 0.858 | -0.004 | 0.087 | 0.946 | 0.083 | 0.088 | 0.821 | 0.162 | 0.088 | 0.553 |
|  | 0.5 | 0.2 | -0.037 | 0.093 | 0.945 | -0.025 | 0.095 | 0.956 | -0.001 | 0.095 | 0.951 | 0.023 | 0.095 | 0.945 | 0.044 | 0.094 | 0.915 |
|  |  | 0.5 | -0.089 | 0.092 | 0.844 | -0.049 | 0.094 | 0.930 | -0.000 | 0.095 | 0.955 | 0.057 | 0.096 | 0.894 | 0.096 | 0.095 | 0.800 |
|  |  | 1 | -0.147 | 0.090 | 0.622 | -0.078 | 0.092 | 0.874 | 0.010 | 0.094 | 0.945 | 0.105 | 0.096 | 0.789 | 0.194 | 0.097 | 0.471 |
| Z – X heterogeneity |  |  |  |  |  |  |  |  |  |  |  |  |  |  |  |  |  |
| Y | $\beta_{XY}$ | $\delta_{VS}$ | $\beta_{ZVX} = -0.5$ | | | $\beta_{ZVX} = -0.25$ | | | $\beta_{ZVX} = 0$ | | | $\beta_{ZVX} = 0.25$ | | | $\beta_{ZVX} = 0.5$ | | |
|  |  |  | Bias | Se | Cov | Bias | Se | Cov | Bias | Se | Cov | Bias | Se | Cov | Bias | Se | Cov |
| Cont | 0 | 0.2 | -0.006 | 0.069 | 0.958 | -0.004 | 0.063 | 0.957 | -0.001 | 0.058 | 0.956 | -0.002 | 0.054 | 0.947 | -0.001 | 0.050 | 0.962 |
|  |  | 0.5 | -0.001 | 0.092 | 0.956 | -0.004 | 0.071 | 0.956 | 0.000 | 0.058 | 0.958 | 0.001 | 0.049 | 0.946 | -0.001 | 0.042 | 0.946 |
|  |  | 1 | -0.017 | 0.160 | 0.966 | -0.008 | 0.084 | 0.961 | -0.003 | 0.056 | 0.958 | 0.001 | 0.043 | 0.951 | 0.000 | 0.035 | 0.958 |
|  | 0.5 | 0.2 | -0.003 | 0.069 | 0.958 | -0.001 | 0.063 | 0.955 | -0.004 | 0.058 | 0.954 | -0.003 | 0.054 | 0.952 | -0.001 | 0.050 | 0.965 |
|  |  | 0.5 | -0.007 | 0.093 | 0.953 | -0.004 | 0.070 | 0.963 | 0.000 | 0.058 | 0.951 | -0.002 | 0.049 | 0.954 | 0.000 | 0.042 | 0.950 |
|  |  | 1 | -0.011 | 0.161 | 0.963 | -0.002 | 0.083 | 0.948 | 0.000 | 0.056 | 0.954 | 0.001 | 0.043 | 0.952 | -0.001 | 0.035 | 0.933 |
| Bin | 0 | 0.2 | -0.002 | 0.104 | 0.949 | 0.002 | 0.095 | 0.945 | 0.001 | 0.087 | 0.943 | 0.000 | 0.081 | 0.948 | 0.000 | 0.075 | 0.963 |
|  |  | 0.5 | -0.003 | 0.137 | 0.958 | -0.001 | 0.106 | 0.952 | -0.003 | 0.087 | 0.955 | -0.001 | 0.073 | 0.946 | 0.001 | 0.063 | 0.940 |
|  |  | 1 | -0.007 | 0.241 | 0.954 | -0.003 | 0.125 | 0.960 | -0.001 | 0.085 | 0.965 | -0.001 | 0.065 | 0.956 | -0.002 | 0.052 | 0.959 |
|  | 0.5 | 0.2 | -0.010 | 0.118 | 0.944 | -0.002 | 0.106 | 0.958 | -0.002 | 0.097 | 0.952 | 0.001 | 0.091 | 0.953 | 0.004 | 0.086 | 0.942 |
|  |  | 0.5 | -0.023 | 0.158 | 0.964 | -0.006 | 0.118 | 0.954 | -0.002 | 0.097 | 0.959 | 0.006 | 0.082 | 0.951 | 0.011 | 0.072 | 0.949 |
|  |  | 1 | -0.070 | 0.277 | 0.974 | -0.020 | 0.141 | 0.951 | 0.001 | 0.095 | 0.954 | 0.011 | 0.073 | 0.942 | 0.013 | 0.060 | 0.962 |

Under *Z*-*X* heterogeneity, the degree of selection bias was small. This is not unexpected for the TSLS method which is known to be robust to misspecification of its first-stage model [7]. The TSRI method does not share this property, but nevertheless the bias in our simulations remained small (see also Supplementary Section 7). Standard errors were affected by selection, but coverage figures were at nominal levels.

Additional simulations are reported in Supplementary Sections 3-5. They suggest that effect modification by *U* (Figure 2c) induces similar bias to that of Table 1, that the bias also occurs in IV analyses with survival outcomes and that IPW can adjust for selection bias if data for the effect modifier are available.

The code to run our simulation study is available via the Github repository: https://github.com/agkatzionis/Selection-bias-and-heterogeneity.

## 5. Discussion

While it is known that selection bias can affect IV analyses, attention in the literature has focused on collider bias. In this work, we have argued that selection bias can also occur due to exposure-outcome effect heterogeneity, if a variable that drives this heterogeneity also affects selection. Although we have attempted to illustrate this using DAGs, we emphasise that heterogeneity is a feature of the outcome model and the causal estimand (e.g. it can be due to interactions between variables affecting the outcome) and can be hard to identify using only DAGs.

Even in the absence of selection bias, IV analyses rely on a homogeneity assumption to estimate the ACE. This assumption states that either the instrument-exposure association or the exposure-outcome effect should be homogeneous; or, if they are both heterogeneous, any heterogeneity in one of these quantities is uncorrelated with heterogeneity in the other [8]. We note that selection bias due to heterogeneity can occur even if this assumption holds at a population level; for example, the assumption was satisfied in our simulation study. In Supplementary Section 6, we present another example where selection bias can induce violations of the homogeneity assumption.

IV analyses are sometimes conducted in a two-sample setting. Selection bias can cause similar issues in two-sample IV analyses as those discussed here, with the additional complication that bias can also occur due to systematic differences in the two samples. (see Supplementary Section 6).

In applied analyses, it is important to investigate selection bias due to heterogeneity and attempt to mitigate it when present. If a variable suspected to affect selection is observed, its status as an effect modifier can be tested in a sensitivity analysis based on the selected sample, by including it in the IV model and assessing evidence of interactions between it and the instrument or exposure. If the effect modifier is unobserved and the exposure is continuous, interactions between it and fitted residuals can be assessed instead. IPW can also be used to adjust for selection bias, as demonstrated in Supplementary Section 5. Another possibility is to implement tests for the IV homogeneity assumption in the selected sample [9], or adapt heterogeneity tests from other settings [10] to IV analyses. In addition, bounding methods and simulation studies can serve as sensitivity analyses. Finally, in applications, subject matter knowledge can be used to assess the existence of effect modifiers.

We hope our work will raise awareness among applied researchers regarding selection bias in IV analyses.

## Data Availability

This study relied on simulated data, not real data. The code used to run the simulations for this manuscript is available via the GitHub repository: https://github.com/agkatzionis/Selection-bias-and-heterogeneity

## Acknowledgements

AG and KH are supported by the UK Medical Research Council and the University of Bristol (MRC-IEU core funding, MC_UU_00032/02). FPH is supported by a research productivity fellowship from the Brazilian National Council for Scientific and Technological Development (grant 303880/2023-6).

