## Supplementary Material for "Selection without Collider Stratification can Bias Instrumental Variable Analyses in the Presence of Causal Effect Heterogeneity"

**1. Collider Bias in IV Analyses**

As our manuscript is mainly concerned with selection bias due to causal effect heterogeneity, we only give a brief description of how collider bias affects IV analyses. We refer to the relevant literature for more details.

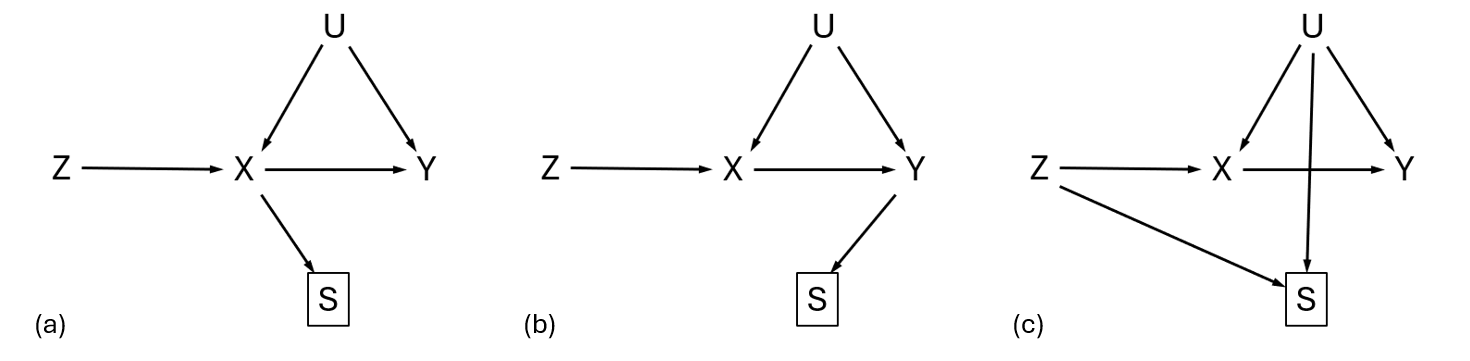

Figure S1: DAGs representing IV analyses where collider bias is induced.

Consider, for example, the DAG of Figure S1a. In this diagram, the exposure affects the selection indicator $S$; the instrument $Z$ and confounder $U$ therefore also affect selection indirectly, via their effect on $X$. Conducting an IV analysis in the selected sample is equivalent to conditioning on $S=1$; this opens the non-causal path $Z\to X\leftarrow U\to Y$ between the instrument and the outcome, inducing bias. This bias occurs regardless of the value of the causal effect and can be induced even under the causal null hypothesis.

A similar bias occurs in the DAG of Figure S1b, where the outcome affects the selection indicator. Again, conditioning on $S=1$ opens the non-causal path $Z\to X\leftarrow U\to Y$ between the instrument and the outcome, inducing bias in IV estimates of the exposure-outcome effect. This bias does not occur under the causal null hypothesis, as in this case $Z$ is not causally upstream of $S$.

Collider bias also occurs in IV analyses if the instrument $Z$ and confounder $U$ affect selection independently of $X$ and $Y$, as shown in Figure S1c. Finally, various combinations of these scenarios may also occur.

**2. More Details about the Simulation Design**

Here, we present the details of the simulation study conducted to illustrate selection bias due to heterogeneity in IV analyses. In our primary simulation study, whose results are reported in the main part of the manuscript, we investigated two scenarios corresponding to the two causal diagrams of Figures 2a and 2b. For the DAG of Figure 2a (effect modification of the $X-Y$ causal effect), we considered a continuous exposure, simulated as

$$X=\beta_{ZX}Z+\beta_{UX}U+\epsilon_{X}$$

The outcome was allowed to be either continuous or binary. Values for the continuous outcome were generated as

$$Y=\beta_{XY}X+\beta_{UX}U+\beta_{VY}V+\beta_{XVY}XV+\epsilon_{Y}$$

while values for the binary outcome were generated as

$$Y\sim Bernoulli\left( \pi_{Y} \right) , logit \pi_{Y}=\beta_{XY}X+\beta_{UX}U+\beta_{VY}V+\beta_{XVY}XV$$

The variables $Z, U,V \epsilon_{X}, \epsilon_{Y}$ were assumed to follow standard normal distributions.

For the DAG of Figure 2b (effect modification of the $Z-X$ association), we generated exposure values as

$$X=\beta_{ZX}Z+\beta_{UX}U+\beta_{VX}V+\beta_{ZVX}ZV+\epsilon_{X}$$

and outcome values as

$$Y=\beta_{XY}X+\beta_{UX}U+\epsilon_{Y}$$

for a continuous outcome and

$$Y\sim Bernoulli\left( \pi_{Y} \right) , logit \pi_{Y}=\beta_{XY}X+\beta_{UX}U$$

for a binary outcome.

Finally, for both causal diagrams, the selection indicator was generated as a logistic random variable

$$S\sim Bernoulli\left( \pi_{S} \right) , logit \pi_{S}=\delta_{0}+ \delta_{VS}V$$

Parameter values were specified as follows. In both simulation scenarios, we set $\beta_{ZX}=0.4$. This value represents an instrument that explains approximately 7.5% of variation in the exposure in simulations with $X-Y$ heterogeneity and 5% of variation in the exposure in simulations with $Z-X$ heterogeneity; in both cases, F statistics were large enough to ensure that our simulation results were not affected by weak instrument bias. The confounder effects were set to $\beta_{UX}=\beta_{UY}=1$, and the value of the causal effect parameter $\beta_{XY}$ was set to be either $0.5$ or $0$. In our first simulation scenario ($X-Y$ heterogeneity, Figure 2a), we set $\beta_{VY}=1$and varied the interaction parameter $\beta_{XVY}$, letting it take values $-0.5, -0.25, 0, 0.25, 0.5$. We also varied the strength of the $V-S$ relationship, to investigate the bias under a weak ($\delta_{VS}=0.2$), moderate ($\delta_{VS}=0.5$) or strong ($\delta_{VS}=1$) selection effect. Similarly, in our second simulation scenario ($Z-X$ heterogeneity, Figure 2b), we set $\beta_{VX}=1$ and varied $\beta_{ZVX}\in\left\{ -0.5, -0.25, 0, 0.25, 0.5 \right\}$ and $\delta_{VS}\in\{0.2, 0.5, 1\}$.

For each scenario and each combination of parameter values, we replicated the simulation 1000 times. In each replication, data were generated for a “population” of $n={10}^{4}$ individuals. A subset of those individuals were then assumed to be selected into the study sample ($S=1$); we set the intercept of the selection model to $\delta_{0}=-0.5$, which resulted in selection probabilities of about 37-41% across all simulations.

The estimand of interest was the ACE. For a continuous outcome, we were interested in the effect on the mean difference scale; this corresponds to the parameter $\beta_{XY}$ in our simulation. To estimate it, we used the two-stage least squares (TSLS) method, with data only on selected individuals. For a binary outcome, we were interested in the effect on the log-odds ratio scale. This is not exactly equal to our parameter $\beta_{XY}$ due to non-collapsibility ($\beta_{XY}$ represents a conditional effect). Therefore, we obtained the true value of the effect by fitting an “oracle” two-stage residual inclusion (TSRI) algorithm to each simulated dataset, using data for all simulated individuals (both those with $S=1$ and those with $S=0$), and averaging the estimates across the 1000 replications. We then obtained sample estimates by fitting the TSRI model only to the selected individuals ($S=1$) in each replication.

As discussed in the main part of the manuscript, our performance measures were the bias in exposure-outcome effect estimates, the average model-based standard error and the empirical coverage of 95% confidence intervals obtained from the TSLS and TSRI methods using only data on selected individuals.

**3. Effect Modification by U**

In this and the next two sections, we extend our original set of simulations by conducting three additional simulation experiments. First, we consider effect modification by an exposure-outcome confounder $U$ and not by an external variable $V$ as previously described. Second, we consider IV analyses with a time-to-event outcome. And third, we investigate whether inverse probability of selection weighting (IPW) can be used to adjust for selection bias due to causal effect heterogeneity.

In our first additional simulation, we considered effect modification by an exposure-outcome confounder $U$, as illustrated in Figure 2c in the main part of the paper. In principle, the confounder $U$ may be an effect modifier for either the instrument-exposure association or the exposure-outcome effect (or both). We considered the two cases separately. Our simulation design was similar to that described earlier, except we did not generate values for the variable $V$ and replaced interaction terms including $V$ with interaction terms including $U$. E.g. in our first scenario ($X-Y$ heterogeneity), values for a continuous outcome were generated according to the model

$$Y=\beta_{XY}X+\beta_{UX}U+\beta_{XUY}XU+\epsilon_{Y}$$

while values for the selection indicator were generated as

$$S\sim Bernoulli\left( \pi_{S} \right) , logit \pi_{S}=\delta_{0}+ \delta_{US}U$$

The results of this simulation are summarized in Table S1 and confirm that selection bias is induced due to effect modification. The magnitude of bias is similar to Table 1, suggesting that, at least in this simple scenario, it does not make much difference whether the effect modifier is an exposure-outcome confounder or another variable. The remarks we made in the main part of our paper also apply to this scenario.

**4. Time-to-event Outcomes**

In our second set of simulations, we considered a scenario with a time-to-event outcome $Y$. As in Table 1 in the main part of the paper, we considered effect modification (by variable $V$) of either the instrument-exposure or the exposure-outcome effect. Data for the outcome were generated according to a Cox-proportional hazards model with a baseline hazard function coming from an Exponential distribution with parameter $\lambda=1$, whence

$$h_{Y}(t)=exp\{\beta_{XY}X+\beta_{UY}U+\beta_{VY}V+\beta_{XVY}XV\}$$

under $X-Y$ heterogeneity and

$$h_{Y}(t)=exp\{\beta_{XY}X+\beta_{UX}U\}$$

under $Z-X$ heterogeneity. For simplicity, we assumed no censoring. We used the “two-stage” method of the R package “ivtools” [1, 2] to perform instrumental variable analysis with survival outcomes.

Table S2 contains the results of this simulation experiment. Under $X-Y$ heterogeneity, the impact of selection bias is similar for the time-to-event outcome as for continuous and binary outcomes. Under $Z-X$ heterogeneity, there is more evidence of bias in Table S2 than in previous tables, and the empirical coverage of 95% confidence intervals is below nominal levels. As with the TSRI method, IV analysis with survival outcomes is not robust to misspecification of the first-stage model, which could explain why more selection bias is induced under $Z-X$ heterogeneity. We note that the empirical coverage of 95% confidence intervals is lower even for $\beta_{XVY}=0$, which corresponds to no effect modification. This issue is unrelated to selection bias and is due to the ivcoxph function’s handling of standard errors: we calculated empirical coverage figures for an oracle IV analysis fitted to both selected and unselected individuals, and the coverage figures for $\beta_{XVY}=0$ were similar to those reported in Table S2.

**5. Inverse Probability Weighting**

IPW is an established method to adjust for selection bias in applications. The use of IPW to deal with selection bias in IV analyses has previously been explored in the context of collider bias [3]. Here, we illustrate that it also can be used to adjust for selection bias due to causal effect heterogeneity.

To do that, we simulated data under the same data generation mechanism as used in our main simulation. For simplicity, we restricted our attention to the case of a continuous outcome. We considered either $X-Y$ or $Z-X$ heterogeneity, using the variable $V$ (and not the confounder $U$) as an effect modifier.

In each replication, weights were calculated by fitting the true missingness model, $logit \pi_{S}=\delta_{0}+ \delta_{VS}V$, to the full simulated dataset. The IV analysis was then implemented as a weighted TSLS fit, with robust standard error estimation using the R package “estimatr”. Results are reported in Table S3.

IPW was able to mitigate selection bias under either $X-Y$ or $Z-X$ heterogeneity and produced estimates with minimal bias and confidence intervals with nominal coverage across all scenarios in this simulation experiment. Therefore, our results confirm that IPW is one potential way to adjust for selection bias in IV analyses, regardless of whether that bias takes the form of collider bias or effect modification.

We emphasize that implementing IPW requires the user to correctly specify a model for the selection process and identify the variables that should be included in that model. In our simulations, this implied that the effect modifier $V$ must be measured for all individuals, both selected and unselected. In practice, this could happen e.g. if $V$ and $X$ are measured when individuals are recruited by the study but the outcome $Y$ is not measured for some individuals due to dropout. However, if the effect modifier is unknown and/or unmeasured, it is not possible to use IPW this way.

**6. Additional Considerations for IV Analyses**

Our simulations suggest that selection bias due to heterogeneity can affect both regression-based analyses and instrumental variable analyses in similar ways. Nevertheless, there are some differences between the two types of analyses that are worth emphasizing.

One such difference is the fact that IV analyses can be biased by heterogeneity in the instrument-exposure relationship, as well as in the exposure-outcome relationship, as we have already explained. In our simulations, we considered the two sources of heterogeneity separately, though in practice, they could co-occur.

Another difference is that IV analyses are often conducted in a two-sample setting, where data for the exposure and outcome may come from different studies. In this case, the default assumption is that the two datasets have been sampled from the same population, although some relaxations of this assumption are possible [4]. In the presence of selection bias, the relationships of the two samples between them, as well as to the target population, must be considered. For example, even in the continuous-outcome scenario of Table 1 where $G-X$ heterogeneity does not bias IV estimates, bias can still be induced in a two-sample IV setting if the two samples systematically differ.

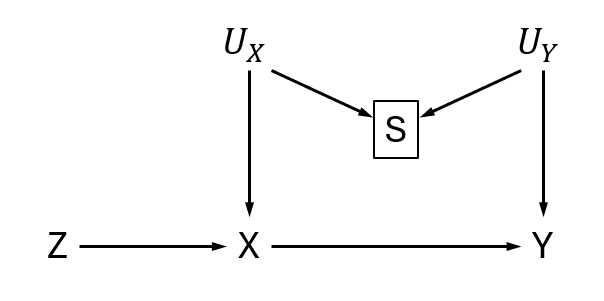

Figure S2: An example DAG where the impact of selection bias on a regression analysis and an IV analysis will be different.

Finally, even if the two types of selection bias affecting regression and IV analyses are similar, the impact of selection bias in the two study designs can be very different in specific applications. For example, consider the causal diagram of Figure S2. Here, $U_{X}$ is an unobserved confounder of $X$ and $S$ that may be an effect modifier of the $Z-X$ relationship, while $U_{Y}$ is an unobserved confounder of $Y$ and $S$ that may be an effect modifier for the $X-Y$ causal effect. In a standard regression analysis (ignoring the instrument $Z$), collider bias will be induced via the path $X\leftarrow U_{X}\to S\leftarrow U_{Y}\to Y$ and bias due to heterogeneity will be induced due to the presence of $U_{Y}$. On the other hand, an IV analysis will not suffer from collider bias in this example, as the path $X\leftarrow U_{X}\to S\leftarrow U_{Y}\to Y$ is structurally a confounding path (it begins with an arrow into $X$ and ends with an arrow into $Y$) and its existence does not violate the IV assumptions (either unconditionally or given $S=1$). However, the IV analysis will be susceptible to selection bias due to heterogeneity in both the instrument-exposure and the exposure-outcome relationships. Effectively, selection induces a violation of the “no simultaneous heterogeneity” assumption, by making the sources of instrument-exposure and exposure-outcome heterogeneity correlated. Given that they have different origins, the biases in the regression and IV analyses may even act in opposite directions. In addition, the regression analysis will be biased by selection even under the causal null hypothesis, while the IV analysis will not be.

**7. Simulations with a Larger Sample Size**

In the main part of our manuscript, we argued that bias can occur for a binary outcome due to heterogeneity in the instrument-exposure association. However, as illustrated in Table 1, this bias is often small. Here, we modify our simulation to provide a better illustration of the existence of bias. Instead of repeatedly simulating small datasets of size $n={10}^{4}$, we generate a single large dataset of size $n={10}^{7}$ for each simulation scenario, to obtain an accurate approximation to the true value of the marginal $X-Y$ causal effect both at a population level and in the selected sample. We then compare the two values to assess whether selection bias occurs. In addition, we consider larger values for some of the simulation parameters was a way to make the bias stronger. For $\delta_{VS}$, we consider the values 1 and 2. For the interaction parameter $\beta_{ZVX}$, we take the values $-1, -0.5, 0, 0.5, 1$. The other parameters are assigned the same values as in the simulations of Table 1.

In Table S4, we report IV estimates and 95% confidence intervals obtained from this simulation experiment. Estimates are reported both at a “population level”, i.e. using all the simulated data without any selection, and in the “selected sample”, i.e. the subset of individuals with $S=1$. As in Table 1, we considered either a normal or a binary outcome. For a normal outcome, our results confirm that no bias is induced in 2SLS estimates: for all values of the parameters $\beta_{XY}, \beta_{ZVX}$ and $\delta_{VS}$, effect estimates from the selected sample are very close to the population-level estimates and the corresponding 95% confidence intervals overlap. For a binary outcome, no bias is induced when $\beta_{XY}=0$, as discussed earlier. However, when $\beta_{XY}=0.5$, we observe deviations between the “population-level” and “sample-based” estimates and the corresponding confidence intervals are disjoint in all cases except when $\beta_{ZVX}=0$ (which indicates no effect modification).

We note that even in this case, the deviations are often small in absolute value, and that this is despite the fact that we have assumed a strong selection effect $\delta_{VS}$ and a strong interaction between $Z$ and $V$. Therefore, for binary outcomes, heterogeneity in the instrument-exposure association may have limited practical consequences compared to other biases (including bias due to exposure-outcome heterogeneity).

| $X-Y$ effect modification by $U$ | | | | | | | | | | | | | | | | | |
| --- | --- | --- | --- | --- | --- | --- | --- | --- | --- | --- | --- | --- | --- | --- | --- | --- | --- |
| $Y$ | $\beta_{XY}$ | $\delta_{US}$ | $\beta_{XUY}= -0.5$ | | | $\beta_{XUY}= -0.25$ | | | $\beta_{XUY}= 0$ | | | $\beta_{XUY}= 0.25$ | | | $\beta_{XUY}= 0.5$ | | |
|  |  |  | Bias | Se | Cov | Bias | Se | Cov | Bias | Se | Cov | Bias | Se | Cov | Bias | Se | Cov |
| Cont | 0 | 0.2 | -0.063 | 0.067 | 0.856 | -0.034 | 0.060 | 0.937 | -0.001 | 0.057 | 0.939 | 0.029 | 0.061 | 0.917 | 0.056 | 0.070 | 0.847 |
|  |  | 0.5 | -0.149 | 0.063 | 0.335 | -0.075 | 0.057 | 0.761 | -0.001 | 0.057 | 0.941 | 0.070 | 0.061 | 0.750 | 0.141 | 0.070 | 0.476 |
|  |  | 1 | -0.254 | 0.057 | 0.002 | -0.127 | 0.053 | 0.327 | -0.002 | 0.054 | 0.948 | 0.125 | 0.059 | 0.419 | 0.250 | 0.068 | 0.067 |
|  | 0.5 | 0.2 | -0.060 | 0.066 | 0.884 | -0.033 | 0.060 | 0.943 | -0.001 | 0.058 | 0.944 | 0.030 | 0.061 | 0.905 | 0.058 | 0.070 | 0.840 |
|  |  | 0.5 | -0.148 | 0.063 | 0.336 | -0.075 | 0.057 | 0.764 | -0.002 | 0.057 | 0.946 | 0.069 | 0.061 | 0.774 | 0.148 | 0.069 | 0.416 |
|  |  | 1 | -0.249 | 0.057 | 0.008 | -0.128 | 0.053 | 0.316 | -0.003 | 0.054 | 0.954 | 0.123 | 0.059 | 0.443 | 0.250 | 0.067 | 0.052 |
| Bin | 0 | 0.2 | -0.039 | 0.086 | 0.924 | -0.017 | 0.087 | 0.957 | 0.000 | 0.089 | 0.947 | 0.019 | 0.089 | 0.929 | 0.032 | 0.089 | 0.932 |
|  |  | 0.5 | -0.085 | 0.084 | 0.839 | -0.051 | 0.086 | 0.907 | 0.003 | 0.089 | 0.951 | 0.048 | 0.090 | 0.916 | 0.088 | 0.090 | 0.827 |
|  |  | 1 | -0.151 | 0.081 | 0.552 | -0.088 | 0.084 | 0.836 | 0.000 | 0.087 | 0.948 | 0.091 | 0.089 | 0.813 | 0.166 | 0.091 | 0.540 |
|  | 0.5 | 0.2 | -0.039 | 0.091 | 0.930 | -0.018 | 0.096 | 0.959 | 0.001 | 0.099 | 0.945 | 0.019 | 0.098 | 0.944 | 0.044 | 0.095 | 0.931 |
|  |  | 0.5 | -0.085 | 0.089 | 0.848 | -0.039 | 0.094 | 0.938 | 0.002 | 0.098 | 0.948 | 0.045 | 0.099 | 0.928 | 0.097 | 0.098 | 0.811 |
|  |  | 1 | -0.147 | 0.085 | 0.603 | -0.063 | 0.092 | 0.881 | 0.004 | 0.097 | 0.947 | 0.079 | 0.099 | 0.875 | 0.184 | 0.099 | 0.552 |
| $Z-X$ effect modification by $U$ | | | | | | | | | | | | | | | | | |
| $Y$ | $\beta_{XY}$ | $\delta_{US}$ | $\beta_{ZUX}= -0.5$ | | | $\beta_{ZUX}= -0.25$ | | | $\beta_{ZUX}= 0$ | | | $\beta_{ZUX}= 0.25$ | | | $\beta_{ZUX}= 0.5$ | | |
|  |  |  | Bias | Se | Cov | Bias | Se | Cov | Bias | Se | Cov | Bias | Se | Cov | Bias | Se | Cov |
| Cont | 0 | 0.2 | 0.000 | 0.068 | 0.948 | -0.003 | 0.063 | 0.955 | -0.002 | 0.058 | 0.961 | -0.002 | 0.054 | 0.956 | -0.003 | 0.050 | 0.950 |
|  |  | 0.5 | -0.003 | 0.090 | 0.949 | -0.004 | 0.069 | 0.953 | 0.002 | 0.056 | 0.951 | -0.003 | 0.048 | 0.947 | -0.001 | 0.041 | 0.961 |
|  |  | 1 | -0.013 | 0.150 | 0.961 | -0.001 | 0.078 | 0.955 | -0.002 | 0.054 | 0.950 | -0.001 | 0.041 | 0.949 | -0.002 | 0.033 | 0.960 |
|  | 0.5 | 0.2 | 0.002 | 0.068 | 0.950 | -0.001 | 0.063 | 0.946 | -0.001 | 0.058 | 0.947 | -0.002 | 0.054 | 0.945 | -0.003 | 0.050 | 0.953 |
|  |  | 0.5 | -0.003 | 0.090 | 0.947 | -0.003 | 0.070 | 0.947 | -0.003 | 0.057 | 0.947 | -0.002 | 0.048 | 0.945 | -0.002 | 0.041 | 0.955 |
|  |  | 1 | -0.015 | 0.151 | 0.955 | -0.007 | 0.079 | 0.956 | 0.000 | 0.054 | 0.963 | -0.002 | 0.041 | 0.953 | 0.000 | 0.033 | 0.951 |
| Bin | 0 | 0.2 | -0.009 | 0.107 | 0.950 | -0.009 | 0.098 | 0.945 | -0.003 | 0.090 | 0.945 | 0.005 | 0.083 | 0.960 | 0.002 | 0.078 | 0.952 |
|  |  | 0.5 | -0.038 | 0.144 | 0.955 | -0.017 | 0.110 | 0.947 | -0.005 | 0.089 | 0.957 | 0.007 | 0.075 | 0.943 | 0.014 | 0.065 | 0.943 |
|  |  | 1 | -0.076 | 0.247 | 0.960 | -0.026 | 0.128 | 0.952 | -0.002 | 0.087 | 0.947 | 0.011 | 0.066 | 0.943 | 0.016 | 0.053 | 0.929 |
|  | 0.5 | 0.2 | -0.009 | 0.119 | 0.941 | -0.007 | 0.107 | 0.953 | 0.002 | 0.099 | 0.949 | 0.006 | 0.092 | 0.956 | -0.002 | 0.087 | 0.960 |
|  |  | 0.5 | -0.015 | 0.159 | 0.939 | -0.015 | 0.121 | 0.950 | -0.001 | 0.098 | 0.954 | 0.009 | 0.083 | 0.959 | 0.007 | 0.073 | 0.950 |
|  |  | 1 | -0.017 | 0.271 | 0.964 | -0.018 | 0.142 | 0.956 | 0.008 | 0.097 | 0.938 | 0.009 | 0.074 | 0.948 | 0.006 | 0.061 | 0.944 |

Table S1. Bias, model-based standard error (“Se”) and empirical coverage (“Cov”) of TSLS/TSRI estimates for various parameter values in a simulation study where the confounder $U$ acts as an effect modifier: $\beta_{XY}$ represents the exposure-outcome causal effect, $\delta_{US}$ the effect of $U$ on selection, and $\beta_{XUY}$ ($\beta_{ZUX}$) the strength of interaction between $X$ and $U$ in their effects on $Y$ (or $Z$ and $U$ in their effects on $X$).

| $X-Y$ heterogeneity - Time-to-event Outcome | | | | | | | | | | | | | | | | | |
| --- | --- | --- | --- | --- | --- | --- | --- | --- | --- | --- | --- | --- | --- | --- | --- | --- | --- |
| $Y$ | $\beta_{XY}$ | $\delta_{VS}$ | $\beta_{XVY}= -0.5$ | | | $\beta_{XVY}= -0.25$ | | | $\beta_{XVY}= 0$ | | | $\beta_{XVY}= 0.25$ | | | $\beta_{XVY}= 0.5$ | | |
|  |  |  | Bias | Se | Cov | Bias | Se | Cov | Bias | Se | Cov | Bias | Se | Cov | Bias | Se | Cov |
| Cox | 0 | 0.2 | -0.036 | 0.040 | 0.838 | -0.019 | 0.041 | 0.904 | 0.001 | 0.042 | 0.934 | 0.018 | 0.043 | 0.921 | 0.028 | 0.044 | 0.869 |
|  |  | 0.5 | -0.083 | 0.040 | 0.460 | -0.044 | 0.040 | 0.786 | 0.001 | 0.042 | 0.930 | 0.040 | 0.043 | 0.809 | 0.070 | 0.045 | 0.634 |
|  |  | 1 | -0.144 | 0.039 | 0.050 | -0.078 | 0.040 | 0.519 | -0.002 | 0.041 | 0.943 | 0.077 | 0.043 | 0.562 | 0.129 | 0.045 | 0.216 |
|  | 0.5 | 0.2 | -0.039 | 0.042 | 0.828 | -0.016 | 0.043 | 0.927 | 0.002 | 0.045 | 0.927 | 0.022 | 0.048 | 0.910 | 0.034 | 0.051 | 0.857 |
|  |  | 0.5 | -0.086 | 0.041 | 0.462 | -0.043 | 0.042 | 0.818 | 0.004 | 0.044 | 0.925 | 0.048 | 0.048 | 0.798 | 0.084 | 0.051 | 0.591 |
|  |  | 1 | -0.146 | 0.040 | 0.045 | -0.073 | 0.041 | 0.574 | 0.012 | 0.043 | 0.930 | 0.097 | 0.047 | 0.446 | 0.165 | 0.052 | 0.142 |
| $Z-X$ heterogeneity - Time-to-event Outcome | | | | | | | | | | | | | | | | | |
| $Y$ | $\beta_{XY}$ | $\delta_{VS}$ | $\beta_{ZVX}= -0.5$ | | | $\beta_{ZVX}= -0.25$ | | | $\beta_{ZVX}= 0$ | | | $\beta_{ZVX}= 0.25$ | | | $\beta_{ZVX}= 0.5$ | | |
|  |  |  | Bias | Se | Cov | Bias | Se | Cov | Bias | Se | Cov | Bias | Se | Cov | Bias | Se | Cov |
| Cox | 0 | 0.2 | 0.010 | 0.050 | 0.932 | 0.001 | 0.046 | 0.939 | -0.002 | 0.042 | 0.925 | 0.002 | 0.039 | 0.921 | 0.010 | 0.037 | 0.905 |
|  |  | 0.5 | 0.033 | 0.067 | 0.906 | 0.006 | 0.051 | 0.935 | -0.001 | 0.042 | 0.935 | 0.005 | 0.035 | 0.926 | 0.016 | 0.031 | 0.887 |
|  |  | 1 | 0.085 | 0.114 | 0.842 | 0.012 | 0.060 | 0.915 | 0.000 | 0.041 | 0.926 | 0.009 | 0.031 | 0.900 | 0.025 | 0.025 | 0.806 |
|  | 0.5 | 0.2 | 0.009 | 0.053 | 0.933 | 0.002 | 0.048 | 0.920 | 0.001 | 0.044 | 0.933 | 0.001 | 0.042 | 0.919 | 0.008 | 0.040 | 0.923 |
|  |  | 0.5 | 0.029 | 0.071 | 0.915 | 0.005 | 0.054 | 0.949 | -0.001 | 0.044 | 0.936 | 0.005 | 0.038 | 0.913 | 0.017 | 0.033 | 0.874 |
|  |  | 1 | 0.082 | 0.123 | 0.873 | 0.012 | 0.063 | 0.921 | 0.003 | 0.043 | 0.925 | 0.010 | 0.033 | 0.906 | 0.024 | 0.027 | 0.835 |

Table S2: Bias, model-based standard error (“Se”) and empirical coverage (“Cov”) of IV estimates for a time-to-event outcome and various parameter values in our simulation study: $\beta_{XY}$ represents the exposure-outcome causal effect, $\delta_{VS}$ the effect of $V$ on selection, $\beta_{XVY}$ the strength of interaction between $X$ and $V$ in their effects on $Y$ and $\beta_{ZVX}$ the strength of interaction between $Z$ and $V$ in their effects on $X$.

| $X-Y$ heterogeneity - Inverse Probability Weighting | | | | | | | | | | | | | | | | | |
| --- | --- | --- | --- | --- | --- | --- | --- | --- | --- | --- | --- | --- | --- | --- | --- | --- | --- |
| $Y$ | $\beta_{XY}$ | $\delta_{VS}$ | $\beta_{XVY}= -0.5$ | | | $\beta_{XVY}= -0.25$ | | | $\beta_{XVY}= 0$ | | | $\beta_{XVY}= 0.25$ | | | $\beta_{XVY}= 0.5$ | | |
|  |  |  | Bias | Se | Cov | Bias | Se | Cov | Bias | Se | Cov | Bias | Se | Cov | Bias | Se | Cov |
| Cox | 0 | 0.2 | -0.003 | 0.080 | 0.958 | -0.003 | 0.074 | 0.951 | 0.001 | 0.071 | 0.954 | -0.002 | 0.073 | 0.956 | 0.001 | 0.077 | 0.953 |
|  |  | 0.5 | -0.004 | 0.087 | 0.950 | 0.002 | 0.079 | 0.949 | -0.002 | 0.076 | 0.954 | -0.003 | 0.076 | 0.950 | -0.001 | 0.080 | 0.952 |
|  |  | 1 | -0.001 | 0.113 | 0.951 | -0.002 | 0.100 | 0.962 | -0.001 | 0.093 | 0.944 | 0.003 | 0.091 | 0.959 | 0.003 | 0.096 | 0.949 |
|  | 0.5 | 0.2 | 0.001 | 0.080 | 0.948 | 0.000 | 0.074 | 0.942 | -0.003 | 0.072 | 0.944 | -0.004 | 0.073 | 0.940 | -0.005 | 0.078 | 0.952 |
|  |  | 0.5 | 0.001 | 0.087 | 0.942 | 0.001 | 0.079 | 0.951 | -0.002 | 0.076 | 0.956 | -0.001 | 0.076 | 0.951 | 0.001 | 0.080 | 0.950 |
|  |  | 1 | -0.006 | 0.112 | 0.950 | -0.006 | 0.100 | 0.960 | 0.001 | 0.093 | 0.951 | 0.002 | 0.091 | 0.937 | -0.005 | 0.096 | 0.936 |
| $Z-X$ heterogeneity - Inverse Probability Weighting | | | | | | | | | | | | | | | | | |
| $Y$ | $\beta_{XY}$ | $\delta_{VS}$ | $\beta_{ZVX}= -0.5$ | | | $\beta_{ZVX}= -0.25$ | | | $\beta_{ZVX}= 0$ | | | $\beta_{ZVX}= 0.25$ | | | $\beta_{ZVX}= 0.5$ | | |
|  |  |  | Bias | Se | Cov | Bias | Se | Cov | Bias | Se | Cov | Bias | Se | Cov | Bias | Se | Cov |
| Cox | 0 | 0.2 | -0.004 | 0.059 | 0.964 | -0.003 | 0.059 | 0.956 | -0.001 | 0.059 | 0.957 | -0.003 | 0.059 | 0.951 | -0.001 | 0.059 | 0.964 |
|  |  | 0.5 | -0.001 | 0.061 | 0.953 | -0.003 | 0.061 | 0.951 | 0.000 | 0.061 | 0.961 | 0.000 | 0.061 | 0.953 | -0.002 | 0.061 | 0.953 |
|  |  | 1 | -0.004 | 0.069 | 0.950 | -0.006 | 0.070 | 0.964 | -0.004 | 0.069 | 0.954 | -0.001 | 0.069 | 0.959 | -0.004 | 0.070 | 0.957 |
|  | 0.5 | 0.2 | -0.002 | 0.059 | 0.958 | -0.001 | 0.059 | 0.951 | -0.004 | 0.058 | 0.956 | -0.003 | 0.058 | 0.958 | -0.002 | 0.058 | 0.963 |
|  |  | 0.5 | -0.003 | 0.061 | 0.954 | -0.003 | 0.060 | 0.958 | -0.001 | 0.060 | 0.958 | -0.003 | 0.061 | 0.951 | -0.002 | 0.061 | 0.956 |
|  |  | 1 | -0.001 | 0.069 | 0.961 | -0.002 | 0.069 | 0.955 | 0.000 | 0.068 | 0.943 | 0.000 | 0.069 | 0.957 | -0.003 | 0.070 | 0.951 |

Table S3: Bias, model-based standard error (“Se”) and empirical coverage (“Cov”) of IPW estimates for a continuous outcome and various parameter values in our simulation study: $\beta_{XY}$ represents the exposure-outcome causal effect, $\delta_{VS}$ the effect of $V$ on selection, $\beta_{XVY}$ the strength of interaction between $X$ and $V$ in their effects on $Y$ and $\beta_{ZVX}$ the strength of interaction between $Z$ and $V$ in their effects on $X$.

| $\beta_{XY}$ | $\delta_{VS}$ | $\beta_{ZVX}$ | Normal Outcome | | | | Binary Outcome | | | |
| --- | --- | --- | --- | --- | --- | --- | --- | --- | --- | --- |
|  |  |  | Population-level | | Selected Sample | | Population-level | | Selected Sample | |
|  |  |  | Est | 95% CI | Est | 95% CI | Est | 95% CI | Est | 95% CI |
| 0 | 1 | -1 | 0.000 | (-0.002, 0.002) | 0.005 | (-0.009, 0.019) | -0.001 | (-0.005, 0.002) | -0.007 | (-0.028, 0.014) |
|  |  | -0.5 | -0.001 | (-0.003, 0.002) | -0.004 | (-0.014, 0.005) | 0.000 | (-0.004, 0.003) | -0.001 | (-0.015, 0.013) |
|  |  | 0 | -0.001 | (-0.003, 0.002) | -0.001 | (-0.004, 0.003) | 0.001 | (-0.002, 0.004) | 0.002 | (-0.003, 0.007) |
|  |  | 0.5 | 0.001 | (-0.002, 0.003) | 0.000 | (-0.002, 0.002) | 0.001 | (-0.002, 0.005) | 0.001 | (-0.003, 0.004) |
|  |  | 1 | 0.001 | (-0.001, 0.003) | 0.001 | (-0.001, 0.002) | 0.000 | (-0.003, 0.003) | 0.001 | (-0.001, 0.004) |
|  | 2 | -1 | 0.001 | (-0.001, 0.003) | -0.002 | (-0.007, 0.002) | -0.001 | (-0.004, 0.002) | 0.002 | (-0.005, 0.009) |
|  |  | -0.5 | -0.001 | (-0.004, 0.001) | -0.020 | (-0.047, 0.008) | 0.001 | (-0.002, 0.005) | 0.011 | (-0.029, 0.050) |
|  |  | 0 | 0.001 | (-0.001, 0.003) | 0.001 | (-0.002, 0.004) | 0.002 | (-0.001, 0.005) | 0.001 | (-0.005, 0.006) |
|  |  | 0.5 | 0.000 | (-0.002, 0.002) | 0.002 | (0.000, 0.004) | 0.000 | (-0.004, 0.003) | 0.000 | (-0.003, 0.002) |
|  |  | 1 | 0.001 | (-0.001, 0.003) | 0.000 | (-0.001, 0.001) | 0.001 | (-0.002, 0.004) | 0.000 | (-0.002, 0.002) |
| 0.5 | 1 | -1 | 0.501 | (0.498, 0.503) | 0.495 | (0.482, 0.509) | 0.456 | (0.452, 0.460) | 0.574 | (0.550, 0.598) |
|  |  | -0.5 | 0.500 | (0.498, 0.503) | 0.500 | (0.491, 0.510) | 0.449 | (0.445, 0.453) | 0.403 | (0.387, 0.419) |
|  |  | 0 | 0.497 | (0.495, 0.500) | 0.496 | (0.493, 0.500) | 0.445 | (0.442, 0.449) | 0.446 | (0.440, 0.452) |
|  |  | 0.5 | 0.500 | (0.498, 0.503) | 0.500 | (0.498, 0.502) | 0.449 | (0.445, 0.453) | 0.459 | (0.455, 0.463) |
|  |  | 1 | 0.499 | (0.497, 0.502) | 0.500 | (0.498, 0.501) | 0.456 | (0.452, 0.460) | 0.469 | (0.466, 0.472) |
|  | 2 | -1 | 0.502 | (0.500, 0.504) | 0.496 | (0.491, 0.500) | 0.460 | (0.456, 0.464) | 0.511 | (0.503, 0.519) |
|  |  | -0.5 | 0.500 | (0.498, 0.502) | 0.484 | (0.457, 0.511) | 0.445 | (0.442, 0.449) | 0.227 | (0.179, 0.274) |
|  |  | 0 | 0.499 | (0.497, 0.501) | 0.500 | (0.497, 0.504) | 0.442 | (0.438, 0.445) | 0.445 | (0.439, 0.451) |
|  |  | 0.5 | 0.500 | (0.498, 0.502) | 0.499 | (0.497, 0.501) | 0.448 | (0.445, 0.452) | 0.468 | (0.465, 0.471) |
|  |  | 1 | 0.500 | (0.498, 0.502) | 0.500 | (0.499, 0.501) | 0.459 | (0.455, 0.463) | 0.475 | (0.473, 0.477) |

Table S4. IV estimates (“Est”) and 95% confidence intervals (“95% CI”) from a single replication of our simulation study with a large sample size ($n={10}^{7}$) and instrument-exposure heterogeneity: $\beta_{XY}$ represents the exposure-outcome causal effect, $\delta_{VS}$ the effect of the effect modifier $V$ on selection, and $\beta_{ZVX}$ the strength of interaction between $Z$ and $V$ in their effects on $X$.
